# Association between hair relaxer use and uterine fibroids among women of reproductive age presenting to a national referral hospital in Kenya: a case control study

**DOI:** 10.1101/2025.09.11.25335603

**Authors:** Mary M. Kithikii, Marshal M. Mweu, Jane W. Muita

**Affiliations:** Department of Public and Global Health, Faculty of Health Sciences, University of Nairobi, Nairobi, Kenya

## Abstract

Uterine fibroids (UFs) precipitate significant morbidity in low and middle income countries, yet evidence on their contextual drivers remains scarce. The objective here was to assess the association between HR use and UFs independent of known risk factors as well as to quantify HR use impact in the population of women of reproductive age. A facility-based case control study was employed to assess the HR-UF association among 264 women presenting to the gynaecologic clinic within Kenyatta National Hospital between 23^rd^ October and 19^th^ December 2024. All cases were prospectively recruited while controls were simple randomly sampled from the clinic’s appointment schedule and frequency-matched to cases by day of presentation. A logistic regression model was used to evaluate the HR-UF association, adjusting for the effect of potential confounders. Thereafter, a population attributable fraction (PAF) (along with its confidence interval) was estimated. The odds of UFs among HR users was approximately five times higher than non-users (OR=5.12, 95% CI: 3.00-8.73, *P*<0.001). This relationship was not confounded by the studied covariates. The PAF estimate for the association was 59.2% (95% CI: 47.2-71.7) – suggesting that about three-fifths of UFs in this population could be prevented if HRs were not used. These findings call for more rigorous enforcement of existing legislations that regulate the supply and use of HRs in the country, with a view to safeguarding consumer health.

## Introduction

Benign gynaecologic conditions (BGC) are leading causes of morbidity globally [1]. They constitute 5.1% of all years lost to disability; surpassing the cumulative loss due to TB, HIV/AIDS and malaria [1]. Of all BGCs, uterine fibroids (UFs) remain the most prevalent pelvic tumours that affect women of reproductive age [2]. UFs, otherwise referred to as leiomyomas/myomas, are defined as benign monoclonal tumours that emerge from the myometrium [3].

The burden of UFs has seen a 74.4% surge in the last 3o years [4]. Globally, there was a 60.2% increase in disability-adjusted life years (DALYs) due to UFs [5]. Black women face the highest burden, with over 80% diagnosed with UFs during their lifetime [6]. Moreover, recurrence rates post-myomectomy are as high as 59% within a span of five years among women of African descent [7]. Notable disparities in UF incidence exist in Sub-Saharan Africa (SSA) – Nigeria, Ghana and Kenya reporting incidence rates of 67% [8], 79% [9] and 10-20% [10] respectively. These variations could be attributable to prolonged delays in seeking health care contributing to underreporting [11].

The use of hair relaxers (HRs) may represent a key UF determinant [12]. HRs are defined as hair-straightening products designed to break and restructure disulphide bonds found within the cortical layer of the hair shaft allowing it to attain and maintain a straight appearance [13]. Use of HRs has been linked to an increase in the risk of hormonally-mediated gynaecologic conditions such as uterine, breast and ovarian cancers [12,14,15]. This is because HRs serve as primary exposure pathways for endocrine-disrupting chemicals (EDCs) [16,17]. Examples of EDCs found in HRs include formaldehyde (a carcinogen), phthalates, parabens, mercury, lead, triclosan, benzophenone and phenols [13,18].

Racial disparities in exposure to EDCs have been documented [19,20]. Studies report that black women experience higher levels of exposure leading to more adverse health outcomes [21]. A large prospective cohort study conducted recently in the United States (US) revealed a higher risk of breast cancer in black than white women ascribable to use of chemical hair straighteners [15]. Studies report that hair products tailored specifically for use by black women contain more EDCs compared to others [22] and that black women outspend other ethnic groups on hair styling products [23]. Endocrine disruption and reproductive harm are linked to exposure to more than one EDC [18]. In SSA, 30-49.2% of black women residing in South Africa [24] and 59% of Kenyan women use HRs [25]. Despite their ubiquitous use, to our knowledge, there are no local/regional studies that have sought to evaluate the association between HR use and UFs. Even fewer studies have attempted to describe the frequency and types of HR products used primarily by women in Africa [15].

Occurrence of UFs is multifactorial in aetiology, with demographic, reproductive, biological and lifestyle factors having been identified [26,27]. Potential lifestyle factors include diet, physical activity, body mass index (BMI), alcohol intake and smoking [27,28]. In particular, alcohol has been shown to exert an oestrogenic effect on the myometrium of the uterus predisposing to UFs [29]. Reproductive factors include nulliparity, contraceptive use and time since last birth [27]. Of note, nulliparous women are at a greater risk of UFs compared to parous women – risk reduction being commensurate with the number of children birthed [30,31]. Demographic factors associated with UFs include ethnicity and age [27,32]. The risk of UFs has been shown to increase with age [33]. Biological risk factors for UFs include family history and age at menarche [31,34]. Early menarche is associated with a heightened risk of UFs [31,35,36].

The objective of this study was to quantify the HR-UF association as well as to estimate the impact of HR use in the population of women of reproductive age independent of known risk factors.

## Materials and methods

### Study setting and design

The study was conducted at Kenyatta National Hospital (KNH) situated in Nairobi County, Kenya. It is a national referral hospital that offers a broad range of medical and surgical services and serves a large catchment population, with approximately 949,000 inpatients and 800,000 outpatients registered annually.

This study employed a facility-based case control study design. The rationale for its use was pegged on its suitability for rare outcomes investigation and ease of recruitment of study participants within the facility. This study conformed to the STROBE guidelines for reporting of case-control studies [37].

### Study population

The study population comprised primary/referral female patients presenting to the KNH gynaecologic outpatient clinic for care over a three-month study period (23^rd^ October-19^th^ December 2024).

### Outcome definition

A case participant was a consenting patient of reproductive age having a confirmed UF diagnosis by ultrasonography. Controls were similarly defined as cases, but without UFs based on ultrasonographic examination. Specifically, they were patients presenting to the same clinic with other gynaecologic conditions not considered related to the primary exposure. As such, patients presenting with adenomyosis, endometriosis, infertility or reproductive cancers (ovarian and uterine) were excluded from the study.

### Sample size and participant recruitment

The requisite sample for the study was determined as described by Kelsey *et al*. [38] for case control studies:

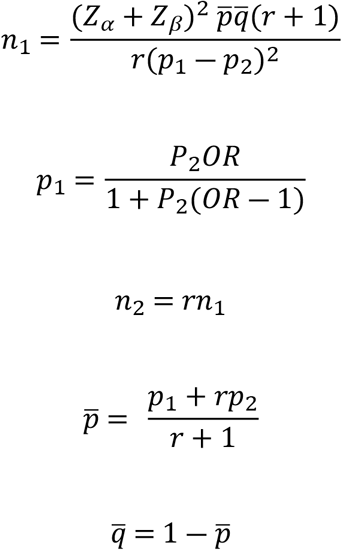

Where: *n*_1_= the number of cases; *n*_2_= the number of controls; *p*_1_= proportion of cases using hair relaxers; *p*_2_= proportion of controls using HRs estimated at 59% [25]. Notably, 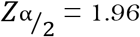 is the value required for a two-sided 95% confidence interval, *Z*_β_= -0.84 is the value of the desired statistical power (80%) and *r*= ratio of controls to cases set at 1. *OR* is the odds ratio for the HR-UF association guesstimated at 2. Considering an anticipated non-response rate of 5%, the sample was calculated to 318 women: 159 cases and 159 controls. Given that the gynaecologic clinic registers up to seven cases daily, to meet the required number, all cases presenting on a particular day were prospectively recruited. Controls were simple randomly sampled from the clinic’s patient registry. They were frequency-matched to cases by day of presentation.

### Study variables

HR use – the study’s primary exposure – related to either their direct use or contact (through inhalation or skin) during application. Other factors recorded included the participant’s sociodemographic (age, ethnicity, marital status, level of education, occupation and income level), lifestyle (body mass index (BMI), physical activity, alcohol intake, biomass fuel exposure and co-morbidities such as diabetes and hypertension), reproductive (contraceptive use and parity) and biological (family history and age at menarche) characteristics. These variables were measured using a semi-structured questionnaire coded within the Open Data Kit (ODK) platform (Table 1). Notably, two research assistants were recruited and trained to assist with the data collection exercise.

**Table 1.**
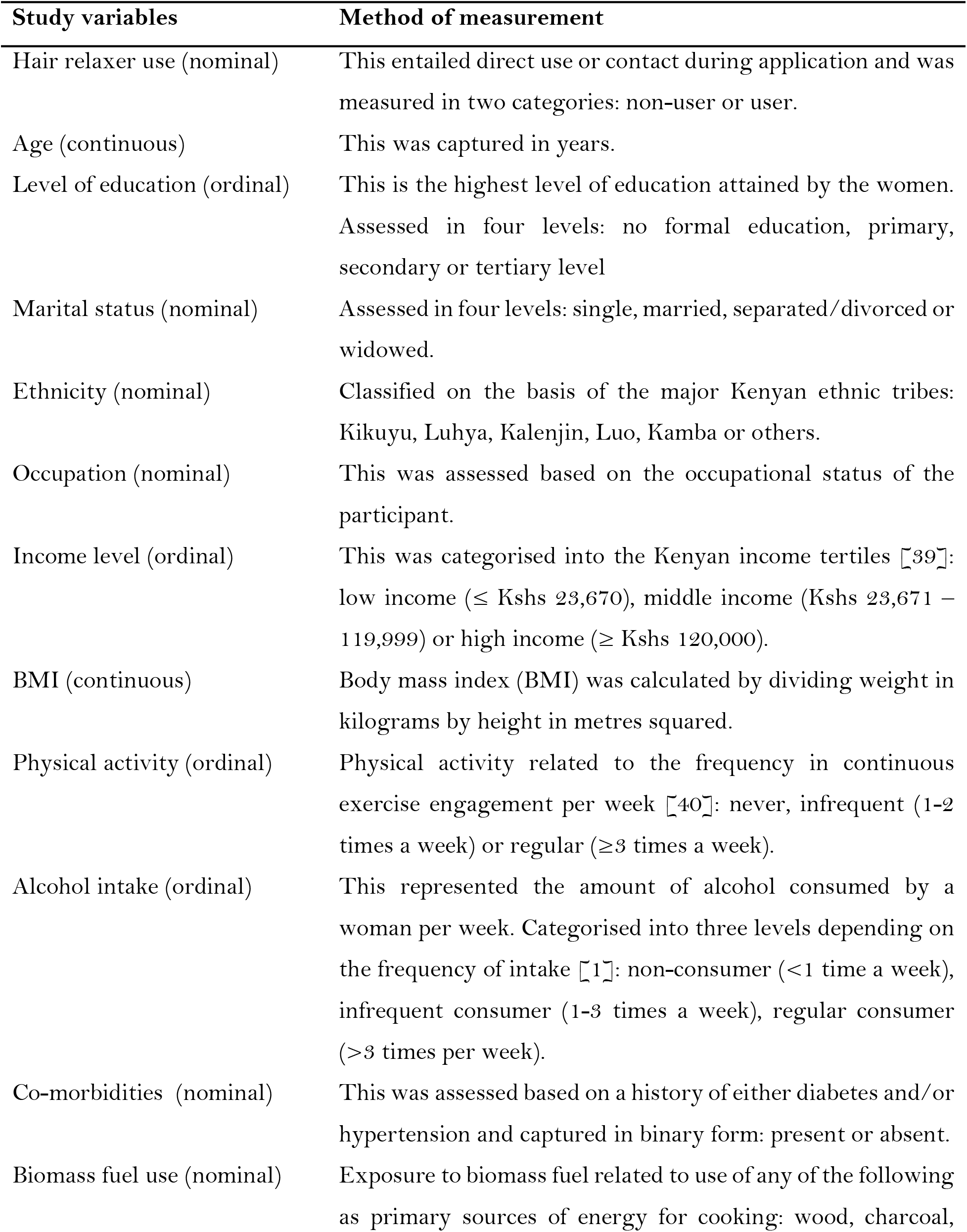
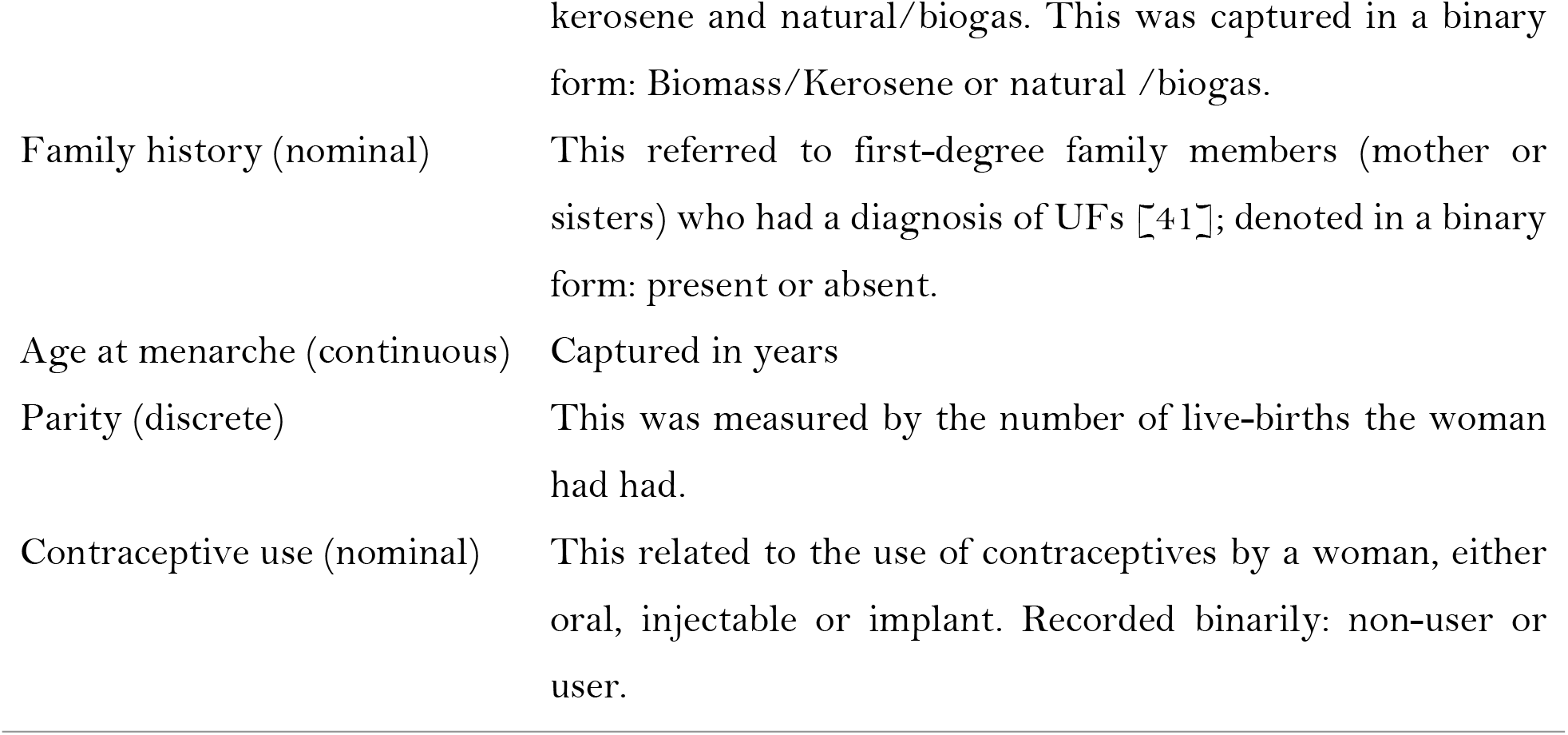
Study variables and their measurements

### Ethical considerations

Approval to conduct the study was granted by the KNH-University of Nairobi Ethics and Research Committee (P487/06/2024) and the National Commission for Science, Technology and Innovation (NACOSTI) (NACOSTI/P/24/41112). Moreover, written informed consent was secured from each individual prior to enrolment in the study.

### Statistical analysis

Initially, the ODK data were exported to R software v4.4.0 for analysis. The R code for these analyses is available as supporting information [42]. For descriptive statistics, qualitative variables were summarised using frequencies and percentages; whilst quantitative variables with medians and ranges.

The crude association between HR use and UF was assessed using a logistic regression model. To evaluate the potential confounding effect of the study covariates on the HR-UF association, all predictors were originally assessed for their unconditional association (amongst HR unexposed women) with UF at a 5% significance level. At this stage, for ease of analysis and owing to a scarcity of observations for some variable categories, recategorisation was effected. Specifically, marital status was regrouped into two categories (single or married), ethnicity was recategorised into five classes (Kikuyu, Luhya, Luo, Kamba and others), level of education was reclassified into three groups (primary, secondary and tertiary levels), income level was dichotomised (low (≤23,670) or high income (>23,670)), alcohol intake converted into a binary variable (no/yes) and BMI was regrouped into four categories (underweight (<18.5 kg/m^2^), normal weight (18.5 – 24.9 kg/m^2^), overweight(25.0 - 29.9 kg/m^2^) or obese (≥ 30.0 kg/m^2^)) [43].

Significant covariates from this step were then screened for an association (amongst controls) with HR use at *P*<0.05. Qualifying variables from this latter stage were considered potential confounders of the HR-UF association and, consequently, added to a multivariable logistic model to adjust for their confounding effect. Importantly, only variables that resulted in a >20% change in the coefficient for HR use following a backward stepwise elimination process were retained in the final multivariable model [44].

The population attributable fraction (PAF) (the proportion of UFs in the population of women of reproductive age that is attributable to HR use), was estimated as specified by Dohoo *et al*. [44]:

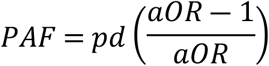

Where: *pd* is the proportion of cases using HRs and *aOR* is the adjusted *OR* for HR use derived from the multivariable model. The estimate’s 95% confidence interval was bootstrapped as described by Ferguson & O’Connell [45].

### Minimisation of errors and biases

To minimise selection bias, recruitment of controls was restricted to patients with conditions not thought to be related to HR use. In order to reduce interviewer bias during elicitation of information from respondents, the research assistants were trained on standardised interviewing techniques. Since differential recall of exposures between cases and controls (hence recall bias) was likely, medical records were referenced to verify reproductive and co-morbidity data. To circumvent data entry errors, the ODK platform was pre-specified with valid values.

## Results

Of the sample of 318 individuals, 264 (159 cases and 105 controls) participated in the study. Among the controls, 24 missed their clinic appointments and 27 declined consent. Descriptive statistics for the study variables are presented in Table 2. In particular, 73.6% (*n*=117) of cases compared with 35.2% (*n*=37) of controls used HRs. The median age of cases was 40 years (range: 20 – 49 years) while that of controls was 31 years (range: 15 – 49 years). Evidently, 49.7% (*n*=79) of cases had a tertiary-level education compared with 59.1% (*n*=62) of controls. Pertaining to contraceptive use, 18.2% (*n*=29) of cases reported using contraceptives in comparison with 39.1% (*n*=41) of controls. The percentage of respondents with a family history of UFs was 25.8% (*n* =41) for cases against 7.6% (*n*=8) for controls.

**Table 2.** Descriptive statistics for the predictors of uterine fibroids among women of reproductive age, Kenyatta National Hospital, Kenya (*n* = 264)

| Variable | Values | Cases<br>$n$ (%) | Controls<br>$n$ (%) |
| --- | --- | --- | --- |
| Marital Status | Single | 65(40.9) | 35(33.3) |
|  | Married | 85(53.5) | 68(64.8) |
|  | Separated/Divorced | 8(5.0) | 2(1.9) |
|  | Widowed | 1(0.6) | 0(0.0) |
| Ethnicity | Kikuyu | 68(43.0) | 40(38.1) |
|  | Luhya | 24(15.2) | 9(8.6) |
|  | Kalenjin | 2(1.3) | 9(8.6) |
|  | Luo | 26(16.5) | 13(12.4) |
|  | Kamba | 17(10.8) | 17(16.2) |
|  | Others | 21(13.3) | 17(16.2) |
| Level of Education | No formal education | 4(2.5) | 0(0.0) |
|  | Primary | 29(18.2) | 14(13.3) |
|  | Secondary | 47(29.6) | 29(27.6) |
|  | Tertiary | 79(49.7) | 62(59.1) |
| Occupation | Unemployed | 20(12.6) | 26(24.8) |
|  | Employed | 139(87.4) | 79(75.2) |
| Income level | Low income | 130(85.5) | 78(75.0) |
|  | Middle Income | 22(14.5) | 25(24.0) |
|  | Upper Income | 0(0.0) | 1(1.0) |
| Physical Activity | Never | 37(23.3) | 34(32.4) |
|  | Infrequent | 48(30.2) | 36(34.3) |
|  | Regular | 74(46.5) | 35(33.3) |
| Co-morbidities | Absent | 113(71.1) | 96(91.4) |
|  | Present | 46(28.9) | 9(8.6) |
| Biomass fuel use | Biomass/Kerosene | 36(22.8) | 20(19.1) |
|  | Natural gas/Biogas | 122(77.2) | 85(81.0) |
| Family history | Absent | 118(74.2) | 97(92.4) |
|  | Present | 41(25.8) | 8(7.6) |
| Contraceptive use | Non-user | 130(81.8) | 64(61.0) |
|  | User | 29(18.2) | 41(39.1) |
| Alcohol intake | Non-consumer | 154(96.9) | 102(97.1) |
|  | Infrequent | 3(1.9) | 2(1.9) |
|  | Regular | 2(1.3) | 1(1.0) |
| HR use | Non-user | 42(26.4) | 68(64.8) |
|  | User | 117(73.6) | 37(35.2) |
|  |  | **Cases(Median[Range])** | **Controls(Median[Range])** |
| Age | - | 40.0(20.0, 49.0) | 31.0(15.0, 49.0) |
| BMI | - | 25.7(13.7, 37.9) | 25.4(16.0, 41.5) |
| Age at menarche | - | 14.0(11.0, 20.0) | 14.0(9.0, 18.0) |
| Parity | - | 2.0(0.0, 6.0) | 1.0(0.0, 8.0) |

A description of the types of hair relaxers, frequency and duration of their use is shown in Table 3. Amongst users, the most commonly applied HRs were TCB (58.4%, *n*=90), Dark & Lovely (35.7%, *n*=55) and Venus (17.5%, *n*=27). A majority of the respondents reported using only one HR product (41.3%, *n*=109). The median duration of HR use among cases was 3 years (Range: 0.5 – 25.0 years), while amongst controls it was 2 years (Range: 0.2 – 20.0 years).

**Table 3.** Types, frequency and duration of hair relaxer use among women of reproductive age, Kenyatta National Hospital, Kenya (*n* = 154)

| Types of HRs |  |  |  |  |
| --- | --- | --- | --- | --- |
| Variable | Values | All women<br>$n$ (%) | Cases<br>$n$ (%) | Controls<br>$n$ (%) |
| Dark & Lovely | Yes | 55(35.7) | 48(41.0) | 7(18.9) |
|  | No | 99(64.3) | 69(59.0) | 30(81.1) |
| TCB | Yes | 90(58.4) | 67(57.3) | 23(62.2) |
|  | No | 64(41.6) | 50(42.7) | 14(37.8) |
| Venus | Yes | 27(17.5) | 22(18.8) | 5(13.5) |
|  | No | 127(82.5) | 95(81.2) | 32(86.5) |
| Soft n' Free | Yes | 14(9.1) | 11(9.4) | 3(8.1) |
|  | No | 140(90.9) | 106(90.6) | 34(91.9) |
| Revlon | Yes | 8(15.2) | 8(6.8) | 0(0.0) |
|  | No | 146(94.8) | 109(93.2) | 37(100.0) |
| Motions | Yes | 0(0.0) | 0(0.0) | 0(0.0) |
|  | No | 154(100.0) | 117(100.0) | 37(100.0) |
| L'Oréal | Yes | 2(1.3) | 2(1.7) | 0(0.0) |
|  | No | 152(98.7) | 115(98.3) | 37(100.0) |
| World of Curls | Yes | 1(0.7) | 1(0.9) | 0(0.0) |
|  | No | 153(99.4) | 116(99.2) | 37(100.0) |
| Nice & Lovely | Yes | 9(5.8) | 5(4.3) | 4(10.8) |
|  | No | 145(94.2) | 112(95.7) | 33(89.2) |
| Mega Growth | Yes | 8(5.2) | 7(6.0) | 1(2.7) |
|  | No | 146(94.8) | 110(94.0) | 36(97.3) |
| Movit | Yes | 9(5.8) | 8(6.8) | 1(2.7) |
|  | No | 145(94.2) | 109(93.2) | 36(97.3) |
| Miracle | Yes | 1(0.7) | 1(0.9) | 0(0.0) |
|  | No | 153(99.4) | 116(99.2) | 37(100.0) |
| Miadi | Yes | 5(3.3) | 3(2.6) | 2(5.4) |
|  | No | 149(96.8) | 114(97.4) | 35(94.6) |

| Variable | Values | <u>Frequency of HR use</u> |  |  |
| --- | --- | --- | --- | --- |
|  |  | All women<br>*n* (%) | Cases<br>*n* (%) | Controls<br>*n* (%) |
| Number of products | 1 | 109(41.3) | 81(50.9) | 28(26.7) |
|  | 2 | 32(12.1) | 23(14.5) | 9(8.6) |
|  | 3 | 4(1.5) | 4(2.5) | 0(0.0) |
|  | 4 | 4(1.5) | 4(2.5) | 0(0.0) |
|  | 5 | 4(1.5) | 4(2.5) | 0(0.0) |
|  | 8 | 1(0.4) | 1(0.6) | 0(0.0) |

| Variable | Values | <u>Duration of HR use</u> |  |  |
| --- | --- | --- | --- | --- |
|  |  | All women<br>Median (Range) | Cases<br>Median (Range) | Controls<br>Median (Range) |
| Duration of HR use (in years) | - | 3.0 (0.2, 25.0) | 3.0 (0.5, 25.0) | 2(0.2, 20.0) |

Results of the crude HR-UF association are displayed in Table 4. The odds of UFs among HR users were approximately five times higher (OR = 5.12; 95% CI: [3.00; 8.73]) than non-users.

**Table 4.**
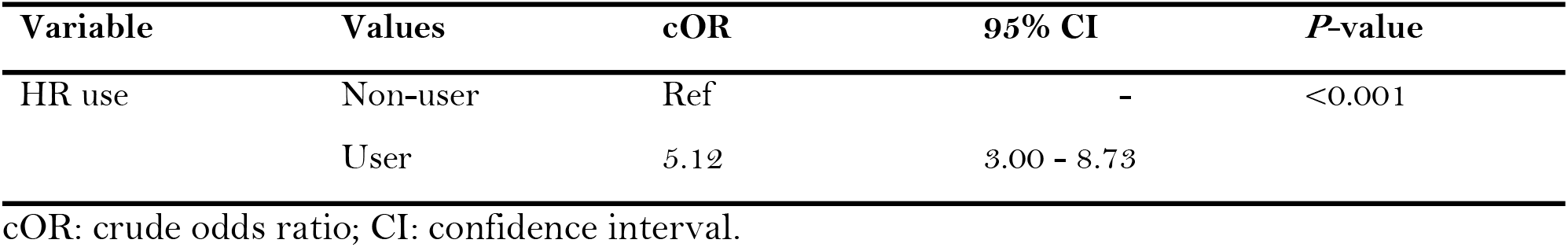
Univariable analysis of the hair relaxer-uterine fibroid association among women of reproductive age, Kenyatta National Hospital, Kenya

| Variable | Values | cOR | 95% CI | P-value |
| --- | --- | --- | --- | --- |
| HR use | Non-user | Ref | - | <0.001 |
|  | User | 5.12 | 3.00 - 8.73 |  |

cOR: crude odds ratio; CI: confidence interval.

Table 5 shows the results of the association between the study variables (amongst HR exposure-negative women) and UF. Notably, age, level of education and contraceptive use were significantly associated with UF and thus qualified for screening with HR use.

**Table 5.** Association between study covariates and uterine fibroids among women of reproductive age, Kenyatta National Hospital, Kenya.

| Variable | Values | OR | 95% CI | P-value |
| --- | --- | --- | --- | --- |
| Age* (in years) | - | 1.13 | 1.06 - 1.20 | <0.001 |
| Marital status | Single | 1.62 | 0.74 - 3.52 | 0.226 |
|  | Married | Ref | - |  |
| Ethnicity | Kikuyu | Ref | - | 0.137 |
|  | Luhya | 3.00 | 0.74 - 12.09 |  |
|  | Luo | 1.54 | 0.51 - 4.71 |  |
|  | Kamba | 0.94 | 0.28 - 3.08 |  |
|  | Others | 0.45 | 0.14 - 1.48 |  |
| Level of Education* | Primary | Ref | - | 0.037 |
|  | Secondary | 0.33 | 0.09-1.22 |  |
|  | Tertiary | 0.22 | 0.07 - 0.73 |  |
| Occupation | Unemployed | Ref | - | 0.306 |
|  | Employed | 1.70 | 0.60 – 4.79 |  |
| Income Level | Low income | 2.08 | 0.74 – 5.79 | 0.146 |
|  | High income | Ref | - |  |
| Family history | Absent | Ref | - | 0.065 |
|  | Present | 2.82 | 0.92 – 8.60 |  |
| Age at menarche | - | 0.97 | 0.78 – 1.22 | 0.814 |
| BMI | Underweight | 0.73 | 0.12 – 4.32 | 0.960 |
|  | Normal | Ref | - |  |
|  | Overweight | 0.97 | 0.37 – 2.55 |  |
|  | Obese | 0.78 | 0.27 – 2.28 |  |
| Physical activity | Never | Ref | - | 0.194 |
|  | Infrequent | 1.48 | 0.57– 3.81 |  |
|  | Regular | 2.38 | 0.92 – 6.16 |  |
| Alcohol intake | No | Ref | - | 0.569 |
|  | Yes | 0.53 | 0.05 – 5.25 |  |
| Biomass fuel use | Biomass/Kerosene | 1.62 | 0.62 – 4.23 | 0.327 |
|  | Natural gas/biogas | Ref | - |  |
| Co-morbidities | Absent | Ref | - | 0.246 |
|  | Present | 2.10 | 0.60 – 7.37 |  |
| Parity |  | 0.97 | 0.75 – 1.25 | 0.794 |
| Contraceptive use* | Non-user | Ref | - | 0.043 |
|  | User | 0.41 | 0.17 – 1.00 |  |

\*Variables eligible for screening for their association with HR use ( $p < 0.05$ ).

**Table 6.**
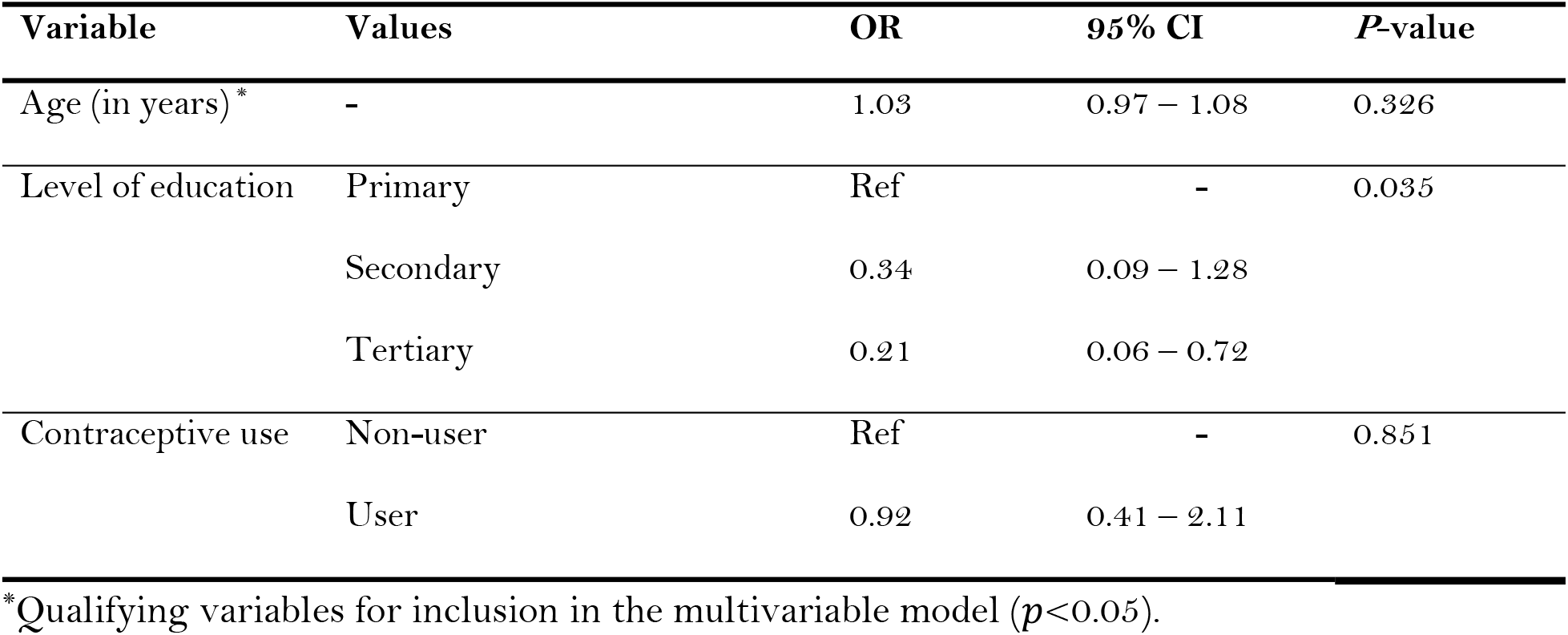
Association between the eligible covariates and hair relaxer use among women of reproductive age, Kenyatta National Hospital, Kenya.

| Variable | Values | OR | 95% CI | P-value |
| --- | --- | --- | --- | --- |
| Age (in years) * | - | 1.03 | 0.97 – 1.08 | 0.326 |
| Level of education | Primary | Ref | - | 0.035 |
|  | Secondary | 0.34 | 0.09 – 1.28 |  |
|  | Tertiary | 0.21 | 0.06 – 0.72 |  |
| Contraceptive use | Non-user | Ref | - | 0.851 |
|  | User | 0.92 | 0.41 – 2.11 |  |

\*Qualifying variables for inclusion in the multivariable model ( $p < 0.05$ ).

Results of the association between qualifying covariates and HR use are presented in Table 6. Only level of education was associated with HR use at the 5% significance level and was thus eligible for assessment of its potential confounding effect on the HR-UF association in the multivariable analysis.

**Table 6:** Multivariable analysis for the association between hair relaxer use and uterine fibroids among women of reproductive age, Kenyatta National Hospital, Kenya.

| Variable | Values | aOR | 95% CI | P-value |
| --- | --- | --- | --- | --- |
| HR use | Non-user | Ref | - | <0.001 |
|  | User | 4.99 | 2.92 – 8.52 |  |
| Level of education | Primary | Ref | - | 0.491 |
|  | Secondary | 0.75 | 0.32 – 1.72 |  |
|  | Tertiary | 0.64 | 0.30 – 1.36 |  |

aOR: adjusted odds ratio.

Table 7 presents the results of the multivariable analysis. Since education level did not confound the HR-UF association (i.e. it did not result in a >20% change in the regression coefficient for HR use) the odds ratio for the unadjusted association was retained and subsequently employed in the estimation of PAF. The PAF estimate for this association was 59.2% (95% CI: 47.2 – 71.7).

## Discussion

In this study setting, women predominantly utilised TCB, Dark & Lovely and Venus HR products. Similarly, in Nakuru, Kenya, Dark & Lovely and TCB were the most desirable brands amongst HR users [2]. These findings underscore the popularity of these products among Kenyan women. The products are available on the market as either lye or no-lye products [46– 48]. Lye HRs contain sodium or potassium hydroxide and are preferred over no-lye HRs due to their greater effectiveness in breaking the disulphide bonds in afro hair shafts resulting in straighter hair [49,50]. Nonetheless, Lye HRs are highly alkaline and have been linked to higher scalp irritations, lesions and burns [50]. No-lye HRs contain calcium hydroxide and guanidine carbonate and are associated with lower irritations or burns [50]. Notwithstanding, a separate study has demonstrated that both lye and no-lye HRs exhibit similar pH and are equally corrosive to the skin [51]. The scalp burns, lesions and irritations caused by these HRs may compromise the skin barrier, potentially increasing dermal absorption of chemicals of concern present in HRs and thereby predisposing to UFs [52,53].

This study demonstrated that a number of women were applying multiple products. This integrated use may synergise their effect, further amplifying the risk of UFs. Frequent use of hair-straightening products has been associated with a two-times higher risk of reproductive cancer [54]. The median duration of HR use for this study population was three years (Range: 0.2, 25.0). Prolonged use of these products could increase exposure to EDCs which could heighten a woman’s risk of UFs. In Ghana, an increasing trend in the long-term usage of no-lye HRs has been observed, with longer duration associated with a higher risk of breast cancer [55]. Comparably, a positive relationship between duration of HR use and incidence of UFs has been noted elsewhere [33].

This study registered a strong association between HR use and UFs even after adjustment for potential confounders – HR users having about five times higher odds of UF than non-users. This reveals that HR use is a strong driver for UFs in this catchment population. This finding is corroborated by results from large prospective studies [12,15,56]. As a case in point, Chang *et al*. [56] demonstrated that women using hair-straightening products had about twice the rate of uterine cancer as non-users. Of note, the HR-UF association in our study yielded a high PAF estimate (59.2%) – signifying that around three-fifths of the UFs in the population could be averted if HRs were not used. This finding calls for dedicated community advocacy campaigns with a view to raising awareness about the potential risks associated with hair-straightening products. A potential mechanism explaining this association is exposure to EDCs commonly found in HRs such as parabens, phthalates, phenols, triclosan, benzophenone and formaldehyde, which are known to influence disease risk [13,18,22,57]. These hormonally-active chemicals perturb endocrine function, potentially contributing to adverse reproductive health outcomes, including UFs [12,22,58]. Indeed, high concentrations of phthalates and paraben metabolites have been recovered from urine samples of fibroid cases [59–62]; connoting a potential causal role [63–66]. Regrettably, consumers may not rely on HR product labels to minimize exposure to EDCs as studies report inconsistencies between listed ingredients and actual measured contents [22,67].

This study is not without its limitations. Despite deliberate efforts to minimise recall bias in the design stage, recall of some exposures was still likely to have been more complete for cases than controls, and this would bias effect estimates away from unity. Population-based (as opposed to hospital-based) controls would have been ideal for this study since they provide a more realistic estimate of the frequency of the exposure (HR use) in the catchment population. However, identifying suitable controls in the population comparable to the hospital cases would have been daunting. Consequently, the reliance on hospital-based controls is likely to have overestimated the frequency of HR use in the population, thus biasing the estimated HR-UF association towards unity.

## Conclusions

A strong association between HR use and UFs was uncovered; HR users having about five times higher odds of UF than non-users. This relationship was not confounded by the studied factors. Accordingly, this association yielded a high PAF – implying that about three-fifths of UFs in the population could be averted devoid of HRs. These findings call for stricter enforcement of existing regulatory frameworks for cosmetic products in the country in order to safeguard consumer health.

## Data Availability

Data are available at Harvard Data verse [Kithikii MM. Replication Data for: Association between hair relaxer use and uterine fibroids among women of reproductive age presenting to a national referral hospital in Kenya: A case control study. Harvard Dataverse 2025. doi:10.7910/DVN/N6YJNG]

https://doi.org/10.7910/DVN/N6YJNG

## Acknowledgments

We are indebted to the hospital’s administration for graciously permitting the conduct of this study.

## References

1. Wijeratne D, Gibson JFE, Fiander A, Rafii-Tabar E, Thakar R. The global burden of disease due to benign gynecological conditions: A call to action. International Journal of Gynecology & Obstetrics. 2023;n/a. doi:10.1002/ijgo.15211

2. Bulun SE. Uterine Fibroids. New England Journal of Medicine. 2013;369: 1344–1355. doi:10.1056/NEJMra1209993

3. Ciavattini A, Di Giuseppe J, Stortoni P, Montik N, Giannubilo SR, Litta P, et al. Uterine Fibroids: Pathogenesis and Interactions with Endometrium and Endomyometrial Junction. Obstetrics and Gynecology International. 2013;2013: e173184. doi:10.1155/2013/173184

4. Lou Z, Huang Y, Li S, Luo Z, Li C, Chu K, et al. Global, regional, and national time trends in incidence, prevalence, years lived with disability for uterine fibroids, 1990-2019: an ageperiod-cohort analysis for the global burden of disease 2019 study. BMC Public Health. 2023;23: 916. doi:10.1186/s12889-023-15765-x

5. Cheng L-C, Li H-Y, Gong Q-Q, Huang C-Y, Zhang C, Yan J-Z. Global, regional, and national burden of uterine fibroids in the last 30 years: Estimates from the 1990 to 2019 Global Burden of Disease Study. Frontiers in Medicine. 2022;9. Available: https://www.frontiersin.org/articles/10.3389/fmed.2022.1003605

6. Baird DD, Dunson DB, Hill MC, Cousins D, Schectman JM. High cumulative incidence of uterine leiomyoma in black and white women: ultrasound evidence. Am J Obstet Gynecol. 2003;188: 100–107. doi:10.1067/mob.2003.99

7. Donnez J, Dolmans M-M. Uterine fibroid management: from the present to the future. Human Reproduction Update. 2016;22: 665–686. doi:10.1093/humupd/dmw023

8. Elugwaraonu O, Okojie AI, Okhia O, Oyadoghan GP. The Incidence of Uterine Fibroid Among Reproductive Age Women: A Five Year Review of Cases at Isth, Irrua, Edo, Nigeria. International Journal of Basic, Applied and Innovative Research. 2013;2: 55–60.

9. Titiloye NA, Duduyemi BM, Asiamah EA, Okai I, Ossei PPS, Konney TO, et al. Total abdominal hysterectomy in a Tertiary Hospital in Kumasi: Indication, Histopathological Findings and Clinicopathological Correlation. Journal of Medical and Biomedical Sciences. 2018;7: 22–28. doi:10.4314/jmbs.v7i1.3

10. U.S. Census Bureau. United States Census Bureau and Population estimates report. 2012 [cited 20 Aug 2012]. Available: Statistics by Country for Uterine Fibroids. Available from: http://www.rightdiagnosis.com/u/uterine_fibroids/stats-country.htm. [Last accessed on 2012 Aug 20].

11. Sabry M, Halder SK, Allah ASA, Roshdy E, Rajaratnam V, Al-Hendy A. Serum vitamin D3 level inversely correlates with uterine fibroid volume in different ethnic groups: a crosssectional observational study. International Journal of Women’s Health. 2013;5: 93–100. doi:10.2147/IJWH.S38800

12. Wise LA, Palmer JR, Reich D, Cozier YC, Rosenberg L. Hair Relaxer Use and Risk of Uterine Leiomyomata in African-American Women. American Journal of Epidemiology. 2012;175: 432–440. doi:10.1093/aje/kwr351

13. DTSCSCPP. Chemicals in Hair Straightening Products Background Document. 2021.

14. Bertrand KA, Delp L, Coogan PF, Cozier YC, Lenzy YM, Rosenberg L, et al. Hair relaxer use and risk of uterine cancer in the Black Women’s Health Study. Environmental Research. 2023;239: 117228. doi:10.1016/j.envres.2023.117228

15. Eberle CE, Sandler DP, Taylor KW, White AJ. Hair dye and chemical straightener use and breast cancer risk in a large US population of black and white women. Int J Cancer. 2020;147: 383–391. doi:10.1002/ijc.32738

16. Braun JM, Just AC, Williams PL, Smith KW, Calafat AM, Hauser R. Personal care product use and urinary phthalate metabolite and paraben concentrations during pregnancy among women from a fertility clinic. J Expo Sci Environ Epidemiol. 2014;24: 459–466. doi:10.1038/jes.2013.69

17. Hsieh C-J, Chang Y-H, Hu A, Chen M-L, Sun C-W, Situmorang RF, et al. Personal care products use and phthalate exposure levels among pregnant women. Science of The Total Environment. 2019;648: 135–143. doi:10.1016/j.scitotenv.2018.08.149

18. Zota AR, Shamasunder B. The environmental injustice of beauty: framing chemical exposures from beauty products as a health disparities concern. American Journal of Obstetrics and Gynecology. 2017;217: 418.e1-418.e6. doi:10.1016/j.ajog.2017.07.020

19. Chan M, Mita C, Bellavia A, Parker M, James-Todd T. Racial/Ethnic Disparities in Pregnancy and Prenatal Exposure to Endocrine-Disrupting Chemicals Commonly Used in Personal Care Products. Curr Envir Health Rpt. 2021;8: 98–112. doi:10.1007/s40572-021-00317-5

20. Schildroth S, Bethea TN, Wesselink AK, Friedman A, Fruh V, Calafat AM, et al. Personal Care Products, Socioeconomic Status, and Endocrine-Disrupting Chemical Mixtures in Black Women. Environ Sci Technol. 2024 [cited 22 Feb 2024]. doi:10.1021/acs.est.3c06440

21. Santaliz Casiano A, Lee A, Teteh D, Madak Erdogan Z, Treviño L. Endocrine-Disrupting Chemicals and Breast Cancer: Disparities in Exposure and Importance of Research Inclusivity. Endocrinology. 2022;163: bqac034. doi:10.1210/endocr/bqac034

22. Helm JS, Nishioka M, Brody JG, Rudel RA, Dodson RE. Measurement of endocrine disrupting and asthma-associated chemicals in hair products used by Black women. Environmental Research. 2018;165: 448–458. doi:10.1016/j.envres.2018.03.030

23. James-Todd T, Connolly L, Preston EV, Quinn MR, Plotan M, Xie Y, et al. Hormonal activity in commonly used Black hair care products: evaluating hormone disruption as a plausible contribution to health disparities. J Expo Sci Environ Epidemiol. 2021;31: 476–486. doi:10.1038/s41370-021-00335-3

24. Khumalo NP, Jessop S, Gumedze F, Ehrlich R. Hairdressing and the prevalence of scalp disease in African adults. British Journal of Dermatology. 2007;157: 981–988. doi:10.1111/j.1365-2133.2007.08146.x

25. Etemesi BA. Impact of hair relaxers in women in Nakuru, Kenya. International Journal of Dermatology. 2007;46: 23–25. doi:10.1111/j.1365-4632.2007.03458.x

26. Pavone D, Clemenza S, Sorbi F, Fambrini M, Petraglia F. Epidemiology and Risk Factors of Uterine Fibroids. Best Practice & Research Clinical Obstetrics & Gynaecology. 2018;46: 3–11. doi:10.1016/j.bpobgyn.2017.09.004

27. Stewart E, Cookson C, Gandolfo R, Schulze-Rath R. Epidemiology of uterine fibroids: a systematic review. BJOG: An International Journal of Obstetrics & Gynaecology. 2017;124: 1501–1512. doi:10.1111/1471-0528.14640

28. He Y, Zeng Q, Dong S, Qin L, Li G, Wang P. Associations between uterine fibroids and lifestyles including diet, physical activity and stress: A case-control study in China. Asia Pacific Journal of Clinical Nutrition. 2020;22: 109–117. doi:10.3316/informit.104887551227284

29. Nagata C, Nakamura K, Oba S, Hayashi M, Takeda N, Yasuda K. Association of intakes of fat, dietary fibre, soya isoflavones and alcohol with uterine fibroids in Japanese women. British Journal of Nutrition. 2009;101: 1427–1431. doi:10.1017/S0007114508083566

30. Wise LA, Palmer JR, Harlow BL, Spiegelman D, Stewart EA, Adams-Campbell LL, et al. Risk of uterine leiomyomata in relation to tobacco, alcohol and caffeine consumption in the Black Women’s Health Study. Human Reproduction. 2004;19: 1746–1754. doi:10.1093/humrep/deh309

31. Wise LA, Laughlin-Tommaso SK. Epidemiology of Uterine Fibroids – From Menarche to Menopause. Clin Obstet Gynecol. 2016;59: 2–24. doi:10.1097/GRF.0000000000000164

32. Murji A, Bedaiwy M, Singh SS, Bougie O. Influence of Ethnicity on Clinical Presentation and Quality of Life in Women With Uterine Fibroids: Results From a Prospective Observational Registry. Journal of Obstetrics and Gynaecology Canada. 2020;42: 726-733.e1. doi:10.1016/j.jogc.2019.10.031

33. Bizjak T, Bečić A. Prevalence and Risk Factors of Uterine Fibroids in North-East Slovenia. Gynecol Obstet. 2016;06. doi:10.4172/2161-0932.1000350

34. Okolo SO, Gentry CC, Perrett CW, Maclean AB. Familial prevalence of uterine fibroids is associated with distinct clinical and molecular features. Human Reproduction. 2005;20: 2321–2324. doi:10.1093/humrep/dei049

35. Schwartz SM. Epidemiology of Uterine Leiomyomata. Clinical Obstetrics and Gynecology. 2001;44: 316.

36. Velez Edwards DR, Baird DD, Hartmann KE. Association of Age at Menarche With Increasing Number of Fibroids in a Cohort of Women Who Underwent Standardized Ultrasound Assessment. American Journal of Epidemiology. 2013;178: 426–433. doi:10.1093/aje/kws585

37. von Elm E, Altman DG, Egger M, Pocock SJ, Gøtzsche PC, Vandenbroucke JP, et al. The Strengthening the Reporting of Observational Studies in Epidemiology (STROBE) statement: guidelines for reporting observational studies. Lancet. 2007;370: 1453–1457. doi:10.1016/S0140-6736(07)61602-X

38. Kelsey JL, Whittemore AS, Evans AS, Thompson WD. Methods in Observational Epidemiology. Oxford University Press; 1996.

39. Kenya National Bureau of Statistics. Economic Survey 2017. 2017. Available: https://www.knbs.or.ke/

40. Yuan He, Qiang Zeng, Sheng-Yong Dong, Li-Qiang Qin, Guo-wei Li, Pei-yu Wang. Associations between Uterine Fibroids and Lifestyles Including Diet, Physical Activity and Stress: A Case-control Study in China. Asia Pacific Journal of Clinical Nutrition. 2013;22. doi:10.6133/apjcn.2013.22.1.07

41. Saldana TM, Moshesh M, Baird DD. Self-reported family history of leiomyoma: not a reliable marker of high risk. Annals of Epidemiology. 2013;23: 286–290. doi:10.1016/j.annepidem.2013.03.003

42. Kithikii MM. Replication Data for: Association between hair relaxer use and uterine fibroids among women of reproductive age presenting to a national referral hospital in Kenya: A case control study. Harvard Dataverse; 2025. doi:10.7910/DVN/N6YJNG

43. WHO. A healthy lifestyle - WHO recommendations. 2010 [cited 5 Jun 2024]. Available: https://www.who.int/europe/news-room/fact-sheets/item/a-healthy-lifestyle---who-recommendations

44. Dohoo IR, Martin SW, Stryhn H. Methods in epidemiologic research. VER Inc. · Charlottetown · Prince Edward Island · Canada; 2012. Available: https://islandscholar.ca/islandora/object/ir%3A8372

45. Ferguson J, O’Connell M. Estimating and displaying population attributable fractions using the R package: graphPAF. Eur J Epidemiol. 2024;39: 715–742. doi:10.1007/s10654-024-01129-1

46. De Sá Dias TC, Baby AR, Kaneko TM, Robles Velasco MV. Relaxing/straightening of Afro-ethnic hair: historical overview. Journal of Cosmetic Dermatology. 2007;6: 2–5. doi:10.1111/j.1473-2165.2007.00294.x

47. Rosenberg L, Boggs DA, Adams-Campbell LL, Palmer JR. Hair Relaxers Not Associated with Breast Cancer Risk: Evidence from the Black Women’s Health Study. Cancer Epidemiology, Biomarkers & Prevention. 2007;16: 1035–1037. doi:10.1158/1055-9965.EPI-06-0946

48. Wickett RR. Permanent waving and straightening of hair. Cutis. 1987;39: 496–497.

49. McMichael AJ. Ethnic hair update: Past and present. Journal of the American Academy of Dermatology. 2003;48: S127–S133. doi:10.1067/mjd.2003.278

50. Richardson V, Agidi AT, Eaddy ER, Davis LS. Ten pearls every dermatologist should know about the appropriate use of relaxers. J of Cosmetic Dermatology. 2017;16: 9–11. doi:10.1111/jocd.12262

51. Sishi VNB, Van Wyk JC, Khumalo NP. The pH of lye and no-lye hair relaxers, including those advertised for children, is at levels that are corrosive to the skin. S Afr Med J. 2019;109: 941–946. doi:10.7196/SAMJ.2019.v109i12.14010

52. Gore AC, Chappell VA, Fenton SE, Flaws JA, Nadal A, Prins GS, et al. EDC-2: The Endocrine Society’s Second Scientific Statement on Endocrine-Disrupting Chemicals. Endocrine Reviews. 2015;36: E1–E150. doi:10.1210/er.2015-1010

53. Yilmaz B, Terekeci H, Sandal S, Kelestimur F. Endocrine disrupting chemicals: exposure, effects on human health, mechanism of action, models for testing and strategies for prevention. Rev Endocr Metab Disord. 2020;21: 127–147. doi:10.1007/s11154-019-09521-z

54. White AJ, Sandler DP, Gaston SA, Jackson CL, O’Brien KM. Use of hair products in relation to ovarian cancer risk. Carcinogenesis. 2021;42: 1189–1195. doi:10.1093/carcin/bgab056

55. Brinton LA, Figueroa JD, Ansong D, Nyarko KM, Wiafe S, Yarney J, et al. Skin lighteners and hair relaxers as risk factors for breast cancer: results from the Ghana breast health study. Carcinogenesis. 2018;39: 571–579. doi:10.1093/carcin/bgy002

56. Chang C-J, O’Brien KM, Keil AP, Gaston SA, Jackson CL, Sandler DP, et al. Use of Straighteners and Other Hair Products and Incident Uterine Cancer. JNCI: Journal of the National Cancer Institute. 2022;114: 1636–1645. doi:10.1093/jnci/djac165

57. James-Todd, Senie R, Terry MB. Racial/Ethnic Differences in Hormonally-Active Hair Product Use: A Plausible Risk Factor for Health Disparities. J Immigrant Minority Health. 2012;14: 506–511. doi:10.1007/s10903-011-9482-5

58. James-Todd T, Connolly L, Preston EV, Quinn MR, Plotan M, Xie Y, et al. Hormonal activity in commonly used Black hair care products: evaluating hormone disruption as aplausible contribution to health disparities. J Expo Sci Environ Epidemiol. 2021;31: 476–486. doi:10.1038/s41370-021-00335-3

59. Branch F, Woodruff TJ, Mitro SD, Zota AR. Vaginal douching and racial/ethnic disparities in phthalates exposures among reproductive-aged women: National Health and Nutrition Examination Survey 2001–2004. Environmental Health. 2015;14: 57. doi:10.1186/s12940-015-0043-6

60. Calafat AM, Ye X, Wong L-Y, Bishop AM, Needham LL. Urinary Concentrations of Four Parabens in the U.S. Population: NHANES 2005–2006. Environmental Health Perspectives. 2010 [cited 1 Apr 2025]. doi:10.1289/ehp.0901560

61. James-Todd TM, Meeker JD, Huang T, Hauser R, Seely EW, Ferguson KK, et al. Racial and ethnic variations in phthalate metabolite concentration changes across full-term pregnancies. J Expo Sci Environ Epidemiol. 2017;27: 160–166. doi:10.1038/jes.2016.2

62. Varshavsky JR, Zota AR, Woodruff TJ. A Novel Method for Calculating Potency-Weighted Cumulative Phthalates Exposure with Implications for Identifying Racial/Ethnic Disparities among U.S. Reproductive-Aged Women in NHANES 2001–2012. Environ Sci Technol. 2016;50: 10616–10624. doi:10.1021/acs.est.6b00522

63. Huang P-C, Li W-F, Liao P-C, Sun C-W, Tsai E-M, Wang S-L. Risk for estrogen-dependent diseases in relation to phthalate exposure and polymorphisms of CYP17A1 and estrogen receptor genes. Environ Sci Pollut Res Int. 2014;21: 13964–13973. doi:10.1007/s11356-014-3260-6

64. Kim JH, Kim SH, Oh YS, Ihm HJ, Chae HD, Kim C-H, et al. In vitro effects of phthalate esters in human myometrial and leiomyoma cells and increased urinary level of phthalate metabolite in women with uterine leiomyoma. Fertility and Sterility. 2017;107: 1061-1069.e1. doi:10.1016/j.fertnstert.2017.01.015

65. Kim YA, Kho Y, Chun KC, Koh JW, Park JW, Bunderson-Schelvan M, et al. Increased Urinary Phthalate Levels in Women with Uterine Leiomyoma: A Case-Control Study. International Journal of Environmental Research and Public Health. 2016;13: 1247. doi:10.3390/ijerph13121247

66. Zota AR, Geller RJ, Calafat AM, Marfori CQ, Baccarelli AA, Moawad GN. Phthalates exposure and uterine fibroid burden among women undergoing surgical treatment for fibroids: a preliminary study. Fertility and Sterility. 2019;111: 112–121. doi:10.1016/j.fertnstert.2018.09.009

67. Dodson RE, Nishioka M, Standley LJ, Perovich LJ, Brody JG, Rudel RA. Endocrine Disruptors and Asthma-Associated Chemicals in Consumer Products. Environmental Health Perspectives. 2012;120: 935–943. doi:10.1289/ehp.1104052

